# Auricular Acupuncture for Erythematotelangiectatic Rosacea: Protocol for a Multicenter, Randomized, Participant- and Assessor-Masked, Sham-Controlled Trial

**DOI:** 10.64898/2026.11.24.26368482

**Authors:** Mingwang Zhang, Xuefu Song, Zhifei Pan, Mei Wan, Zhiqiang Song, Jian Li

**Author notes:** Corresponding author: Correspondence to Jian Li and Zhiqiang Song. The first corresponding author is Jian Li No. 30, Gaotanyan Street, Shapingba District, Chongqing, China.

## Abstract

**Background:** Persistent facial erythema and episodic flushing are prominent features of erythematotelangiectatic rosacea (ETR). The efficacy of auricular acupuncture (AA) for ETR is uncertain.

**Methods:** This multicenter, randomized, participant- and assessor-masked, sham- controlled superiority trial was designed to enroll 108 adults aged 18 to 50 years with ETR and a baseline Clinician Erythema Assessment (CEA) score of at least 2 at 5 hospitals in China. Participants were allocated 1:1 to AA with semipermanent intradermal needles or visually matched, same-site nonpenetrating sham stimulation. Four auricular points (Shenmen, Sanjiao, Endocrine, and Heart) were treated on alternating ears every 3 days for 6 weeks, with self-pressure 3 times daily. Follow-up was scheduled through week 10. The primary outcome was clinician-rated erythema (CEA score) at week 6.

Secondary outcomes comprised other clinician-rated erythema assessments, patient-rated redness, flushing, migraine symptoms, sleep quality, depressive and anxiety symptoms, and adverse events. The primary treatment contrast was planned using baseline-adjusted analysis of covariance with multiple imputation for missing week 6 outcomes. Secondary longitudinal outcomes were planned for exploratory mixed-model analyses.

**Discussion:** The trial was designed to assess the incremental efficacy and tolerability of penetrating auricular needling compared with a matched nonpenetrating procedure.

Same-site pressure in both groups may have physiological effects, which should be considered when interpreting the results.

**Trial registration:** International Traditional Medicine Clinical Trial Registry, ITMCTR2024000324.

## 1 Introduction

### 1.1 Background and Scientific Rationale

Rosacea is a chronic inflammatory disorder of the central face characterized by recurrent flushing, persistent erythema, telangiectasia, inflammatory lesions, and, in some patients, ocular or phymatous involvement. A systematic review estimated that rosacea affects 5.46% of adults worldwide ^1^. ETR is distinguished by persistent centrofacial erythema with episodic intensification and visible telangiectasia, making facial redness the dominant clinical problem in this population.

The burden of ETR extends beyond visible skin changes. Rosacea is associated with impaired quality of life and increased risks of depression, anxiety and sleep disturbance, and epidemiologic evidence also supports an association with migraine ^2,3^. Available treatments for persistent erythema include topical alpha-adrenergic agonists and vascular laser or light-based procedures. Systemic agents such as beta-blockers are used off label in selected patients. These approaches may improve individual features, but responses are variable, recurrence is common, and treatment may not address the neurogenic or psychosocial symptoms that accompany ETR ^4^. A treatment that can improve facial erythema without adding sustained systemic drug exposure would therefore address an important clinical need.

Current models of rosacea pathophysiology implicate interacting neurovascular and immune processes rather than isolated vascular dilatation. Environmental and emotional triggers can activate cutaneous sensory pathways, promote release of vasoactive neuropeptides, and amplify innate immune signaling, thereby linking flushing, persistent erythema, and inflammation ^5^. This neuroimmune framework provides a rationale for testing neuromodulatory interventions, while recognizing that clinical efficacy cannot be inferred from mechanistic plausibility alone.

AA uses needles placed at defined points on the auricular pavilion and has developed within both traditional East Asian and modern European auricular treatment systems ^6,7^. The external ear has heterogeneous sensory innervation that includes the auricular branch of the vagus nerve, particularly within the conchal region ^8,9^. Experimental studies show that vagal stimulation can attenuate proinflammatory cytokine responses, and preclinical auricular stimulation studies support engagement of vagally mediated anti-inflammatory pathways ^10,11^. These findings make auricular neuromodulation a biologically plausible approach for ETR, but they do not establish that needling the selected auricular points will improve rosacea. Randomized evidence in depression indicates that repeated semipermanent auricular needling can be delivered in outpatient settings, although local pain is common and may lead to treatment discontinuation ^12^. At the time this protocol was developed, the evidence reviewed for the study did not include a randomized trial of AA for ETR. A rigorously masked, sham-controlled trial was therefore required to estimate its clinical effects and harms.

### 1.2 Rationale for the Comparator

A nonpenetrating sham applied at the same auricular points was selected to match the procedural and contextual components of AA while estimating the incremental effect of penetrating needling. Because auricular pressure may itself produce sensory stimulation, the sham should not be regarded as physiologically inert. Device preparation and application are specified in Section 4.7, and masking procedures are specified in Section 6.4.

### 1.3 Benefit-Risk Assessment

Participants assigned to AA may experience improvement in persistent erythema, flushing, or associated patient-reported symptoms, but individual benefit is uncertain. Participants in both groups may benefit indirectly from structured dermatologic assessment, repeated follow-up, and prompt identification of clinical deterioration or AEs. The study may also provide evidence on a low-cost, nonpharmacologic intervention that could be delivered in outpatient care if efficacy and acceptable tolerability are demonstrated.

Foreseeable intervention-related risks included localized pain or intolerance, adhesive allergy, bleeding, and auricular infection. Participants were required to attend repeated treatment visits, wear the devices for 3 days, press each point 3 times daily, and complete clinical and questionnaire assessments. Risk mitigation included sterile single-use active devices, skin disinfection, trained practitioners, standardized procedures, auricular inspection at treatment visits, and active adverse event monitoring. For sham treatment, practitioners disinfected the trimmed contact surface with povidone-iodine before application without repackaging.

Treatment was discontinued if an allergic reaction, infection, or other clinically important event made continuation inappropriate.

### 1.4 Objectives and Hypotheses

**Primary objective.** To determine whether AA reduces clinician-rated facial erythema at week 6 compared with SS in ETR, as measured by the CEA score.

**Secondary objectives.** To assess between-group differences in CEA scores at weeks 2, 4, and 10 and in PSA, GFSS, VAS, PSQI, HAMD, and HAMA scores at weeks 2, 4, 6, and 10. Treatment safety and adverse events will also be evaluated throughout follow-up.

**Primary hypothesis.** Participants assigned to AA will have a lower mean CEA score at week 6 than participants assigned to SS after adjustment for baseline CEA score.

**Secondary hypotheses.** AA will be associated with more favorable trajectories of clinician-rated and participant-reported erythema and related symptoms through week 10. Safety outcomes will be summarized descriptively, without a hypothesis of equivalence or noninferiority. Secondary efficacy analyses will be exploratory because no multiplicity adjustment is planned.

## 2 Patient and Public Involvement and Trial Design

### 2.1 Patient and Public Involvement

Patients and members of the public did not participate in developing the research question or designing the trial, intervention, outcomes, recruitment, conduct, oversight, or dissemination plan. Trial participants will provide clinician-rated and patient-reported outcome data as part of study participation, but this does not constitute patient and public involvement in trial design or governance.

### 2.2 Trial Design

This is a multicenter, parallel-group, randomized, participant- and assessor-masked, sham- controlled superiority trial conducted at 5 hospital-based clinical sites in China. Eligible adults with ETR will be allocated in a 1:1 ratio to receive either AA or SS. The assigned intervention will be administered for 6 weeks, with outcome assessments at baseline and weeks 2, 4, 6, and 10. The primary comparison will estimate the effect of assignment to AA versus SS on the CEA score at week 6. The week 10 assessment will evaluate whether any effects observed at the end of treatment persist during the 4-week post-treatment follow-up.

Participants, dermatologists, psychiatrists, and the trial statistician will remain masked to treatment allocation. Treating acupuncturists will be aware of allocation but will not assess outcomes or perform statistical analyses. The 2 devices will appear identical, and treatment procedures will be matched between groups. Allocation will be generated and concealed through the central electronic data capture (EDC) system. Further masking procedures are described in Section 6.

### 2.3 Study Setting and Participating Sites

The trial will be conducted in outpatient dermatology settings at the following 5 hospitals in China: the Department of Dermatology, The First Affiliated Hospital of Army Medical University; the Department of Dermatology, General Hospital of Western Theater Command of the People’s Liberation Army; the Department of Dermatology, The 920th Hospital of the Joint Logistic Support Force of the People’s Liberation Army; the Department of Dermatology, the 958 Jiangbei Branch of the First Affiliated Hospital of Army Medical University; and the Department of Dermatology, Shapingba District Hospital of Traditional Chinese Medicine, Chongqing. The First Affiliated Hospital of Army Medical University will serve as the coordinating center and will oversee central randomization, data management, and cross-site administrative coordination.

Before enrollment, each site was required to have ethics approval, access to eligible adults with ETR, qualified dermatologists and acupuncture practitioners, suitable treatment and follow-up facilities, and the common protocol, Case Report Forms (CRFs) and intervention supplies.

## 3 Participants

### 3.1 Target Population

Adults aged 18 to 50 years, of any sex, will be recruited from the outpatient services of the participating hospitals. The target population comprises patients with ETR diagnosed in accordance with the global ROSCO ^13^ recommendations and with clinically appreciable persistent facial erythema, defined as a baseline CEA score of 2 or greater independently confirmed by 2 dermatologists.

Participants must be willing and able to attend the assigned treatment sessions and follow-up visits, complete the prespecified assessments, and cooperate with efficacy and AE monitoring. These criteria define a clinically relevant population in whom change in persistent facial erythema can be measured while maintaining the feasibility and safety of the 10-week trial schedule.

### 3.2 Inclusion Criteria

1. Age 18 to 50 years, inclusive, of any sex.
2. A diagnosis of ETR according to the ROSCO recommendations and a baseline CEA score of 2 or greater, independently confirmed by 2 dermatologists.
3. Willingness and ability to comply with the assigned treatment and follow-up schedule, complete study assessments, and cooperate with efficacy and AE monitoring.
4. Provision of written informed consent before any study-specific procedures.

### 3.3 Exclusion Criteria

1. Papulopustular, ocular, or phymatous rosacea.
2. Participation in an investigational drug study within 1 month before enrollment.
3. Current use of medication that may affect rosacea, including carvedilol, gabapentin, duloxetine, minocycline hydrochloride, or isotretinoin.
4. Bradycardia, severe cardiac, hepatic, pulmonary, renal, or hematopoietic disease, or another condition that, in the investigator’s judgment, makes trial participation inappropriate.
5. Malignant disease, systemic failure, or an immunocompromised state.
6. Facial acne, atopic dermatitis, steroid-dependent dermatitis, or another facial dermatosis that could interfere with assessment of erythema.
7. A personal or family history of epilepsy, a tic disorder, or another neuropsychiatric condition that would prevent cooperation with trial procedures.
8. A contraindication to acupuncture, fear of needling, or inability to tolerate pain.
9. An auricular abscess, infection, ulceration, edema, or scar, or known allergy to the study- device adhesive tape used with the study device.
10. Auricular cartilage disease, including polychondritis or auricular nodules.

### 3.4 Eligibility of Sites and Intervention Providers

Site requirements are specified in Section 2.3. All 5 participating centers have designated AA treatment areas and previous experience providing clinical acupuncture services. Practitioner qualifications and standardized intervention procedures are specified in Section 4.6. Masking and the separation of intervention delivery from outcome assessment are specified in Section 6.4.

### 3.5 Recruitment and Screening

Potential participants will be identified during routine outpatient consultations at the 5 participating hospitals. Site dermatologists will conduct an initial clinical prescreening for an ETR phenotype and apparent eligibility. Written informed consent will be obtained before any study-specific screening procedure is performed.

After consent, screening will include demographic and medical information, medication and rosacea history, examination of the face and auricles, and assessment of all inclusion and exclusion criteria. Two dermatologists will independently rate baseline CEA. Only participants with confirmed eligibility will proceed to enrollment and central randomization in the EDC system.

### 3.6 Informed Consent Process

Before any study-specific procedure, a site investigator will explain the study objectives, procedures, random assignment, and masking. The explanation will also cover the expected time commitment, foreseeable risks and potential benefits, treatment alternatives, confidentiality, and voluntary participation. Potential participants will have sufficient time to consider the study and ask questions before signing the written consent form.

A decision not to participate, or a later decision to withdraw, will not result in discrimination or retaliation and will not affect the participant’s usual medical care or legal rights. The signed consent form will be retained in accordance with ethics and institutional requirements.

Participant privacy and the confidentiality of identifiable information will be protected throughout the trial.

### 3.7 Withdrawal, Intervention Discontinuation, and Loss to Follow-up

**Withdrawal of consent.** Participants may withdraw consent at any time without penalty. The study team will, if the participant is willing, clarify whether the withdrawal applies to the assigned intervention only, to further study visits and data collection, or to all remaining trial participation, and will document the date, scope, and reason when provided.

**Intervention discontinuation.** Criteria for discontinuing the assigned intervention and the required clinical actions are specified in Section 4.8.

**Follow-up after intervention discontinuation.** Discontinuation of the assigned intervention does not by itself constitute withdrawal from the trial. Unless the participant withdraws permission for further follow-up, the study team will seek to complete the scheduled outcome and safety assessments through week 10. The participant will remain in the originally assigned group for the ITT analysis, and data collected before discontinuation will be retained and used as permitted by the informed consent and applicable requirements.

**Loss to follow-up.** Missed visits and attempts to restore contact will be managed and documented as specified in Section 7.3. No new data will be collected after complete withdrawal of consent. Previously collected data will be handled according to the consent and applicable requirements.

## 4 Interventions and Comparator

### 4.1 Intervention Overview

Both randomized groups will receive a fixed unilateral intervention at 4 auricular points in the following order: Shenmen (TF4), Sanjiao (CO17), Neifenmi (Endocrine, CO18), and Xin (Heart, CO15). These standard codes identify the triangular fossa (TF) and concha (CO) according to GB/T 13734-2008 (Nomenclature and Location of Auricular Points). In the AA group, the intact 2.0-mm sterile press needles will penetrate the selected points. In the SS group, visually matched nonpenetrating devices will be applied at the same points. Point selection will not be individualized. Device construction is described in Section 4.7.

Both groups will receive the same skin preparation, number of devices, unilateral application, 3- day retention period, alternating-ear regimen, 6-week treatment course, participant self-pressing instructions, contact with the intervention provider, and safety surveillance. No study intervention will be delivered during the post-treatment follow-up from week 6 through week 10.

The intervention and comparator are specified below in accordance with the STRICTA reporting framework ^14^.

### 4.2 AA Rationale

The intervention is a standardized form of traditional Chinese AA using semipermanent intradermal needles. The fixed prescription was selected before trial initiation through discussion by the research team, informed by traditional auricular theory, published auricular therapy literature, and the proposed neuroimmune and neurovascular mechanisms of ETR. No point will be added, omitted, or substituted according to an individual participant’s symptom pattern.

The biomedical rationale is that the auricle has heterogeneous sensory innervation, including the auricular branch of the vagus nerve, and that auricular stimulation may engage autonomic and anti-inflammatory pathways. Contemporary randomized trials have also shown that standardized auricular needling with multiday retention and participant self-stimulation can be delivered reproducibly in outpatient settings, although the indications and point prescriptions differ from the present trial ^12,15^.

### 4.3 Needling Details

Four insertions will be made at each treatment application on 1 ear. Practitioners will identify the fixed auricular points using anatomic landmarks according to GB/T 13734-2008. The application order is Shenmen (TF4), Sanjiao (CO17), Neifenmi (Endocrine, CO18), and Xin (Heart, CO15). Shenmen is located in the upper posterior triangular fossa. Sanjiao is posteroinferior to the external acoustic meatus, between the Lung and Endocrine regions. Endocrine is located within the intertragic notch at the floor of the cavum conchae, and Heart is in the central depression of the cavum conchae. The same points and sequence will be used in both groups at every application (Figure 1).

**Figure 1.**
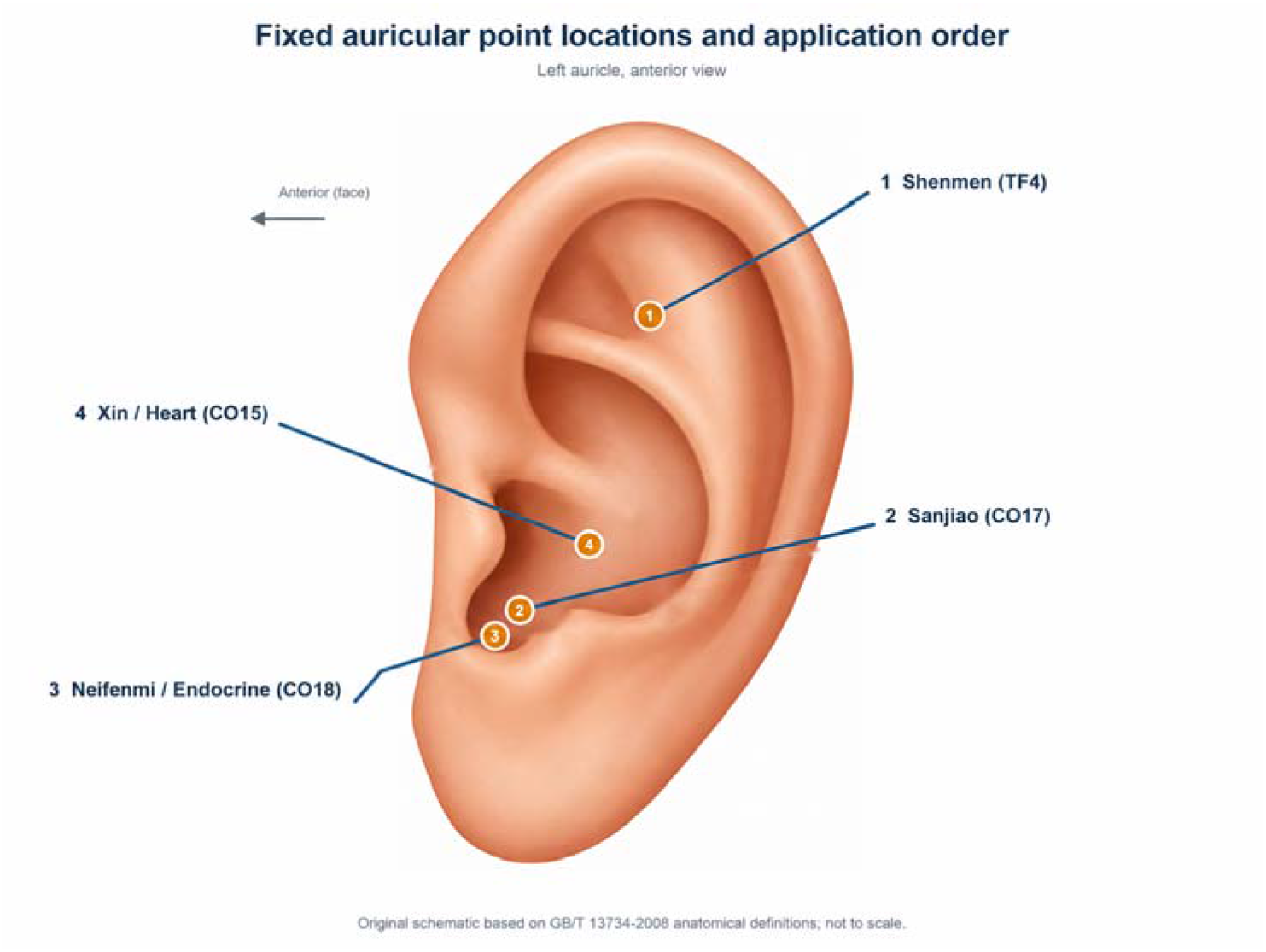
Standardized Auricular Point Locations and Application Order. An anterior view of the left auricle identifies the points in the fixed application sequence: 1, Shenmen (TF4); 2, Sanjiao (CO17); 3, Neifenmi (Endocrine, CO18); and 4, Xin (Heart, CO15). Locations are based on GB/T 13734-2008.

Participants will remain seated during device placement. The auricular skin will be cleaned with a moist swab, disinfected with Aner iodine antiseptic, and dried. For AA, a JIAJIAN sterile press needle (SPN; 0.22-mm diameter and 2.0-mm needle length; Wuxi Jiajian Medical Instrument Co, Ltd) will be inserted approximately perpendicular to the auricular surface to a depth of 2.0 mm and secured with its integral adhesive. According to the manufacturer, the single-use, ethylene-oxide-sterilized device contains 06Cr19Ni10 austenitic stainless steel and carries Jiangsu medical device registration certificate 20152200876 (Su Xie Zhu Zhun). No electrical or instrument-assisted stimulation will be used. The devices are illustrated in Figure 2.

**Figure 2.**
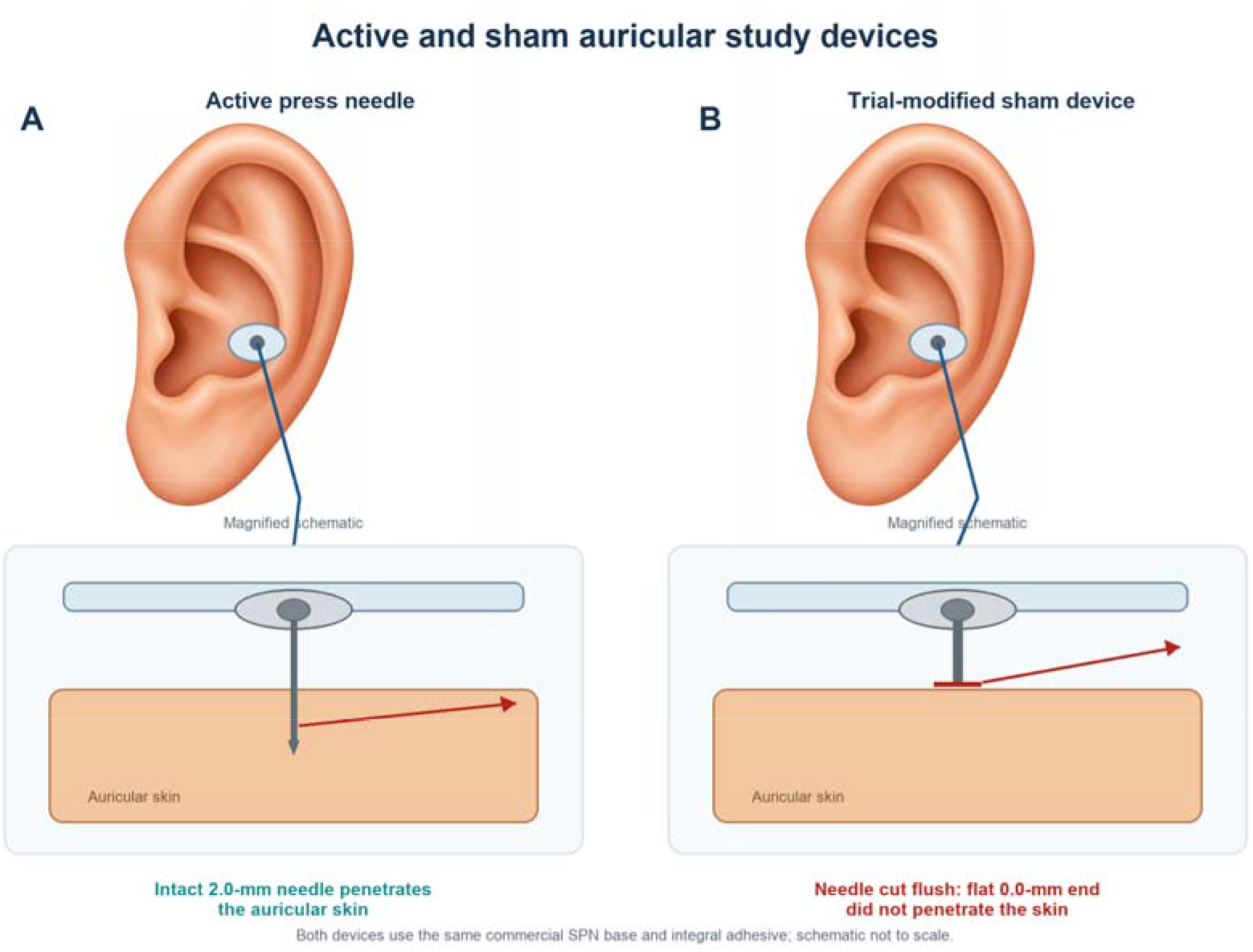
Active and Sham Study Devices. (A) Commercial JIAJIAN SPN (0.22-mm diameter, 2.0-mm needle length) penetrating the auricular skin. (B) Sham device prepared from the same product by cutting the needle flush to form a flat, nonpenetrating contact surface. After trimming, the contact surface was disinfected with povidone-iodine and applied without repackaging. The adhesive and outward appearance were unchanged.

Immediately after placement, the acupuncturist will manually press each device to an intensity tolerated by the participant. The response sought is a de qi-like sensation, which may include soreness, numbness, heaviness, distension, or pain, together with visible auricular erythema and subjective warmth. Each device will remain in place for 3 days. Device diameter, length, lot information, treated ear, points used, and any procedural deviation will be recorded in the intervention record.

### 4.4 Treatment Regimen

Treatment will begin after randomization and will continue for 6 weeks. All participants will receive the first application on the left ear. A new set of 4 devices will be applied every 3 days, for 14 planned applications: odd-numbered applications on the left ear and even-numbered applications on the right ear. At each return visit, the acupuncturist will remove the devices retained from the preceding 3-day period before applying the next set to the contralateral ear.

The number of applications, device retention, point prescription, and ear alternation will be identical in both groups. Dates of application and removal and the treated ear will be recorded. Missed or incomplete applications will be documented as protocol deviations. The intervention will not be extended beyond the prespecified 6 weeks to replace missed applications.

### 4.5 Other Components, Setting, and Participant Instructions

Applications will be performed in designated outpatient treatment areas by qualified acupuncture practitioners. Clinical outcome assessments will be conducted separately by masked dermatologists. Participants will not be shown the device or application process. The procedural sequence, neutral explanation, application time, practitioner contact, and safety questions will be matched between groups.

Participants in both groups will be verbally instructed to press each of the 4 applied points 3 times daily, once in the morning, once at midday, and once in the evening. Each point will be pressed until the auricle becomes flushed and warm. Pressure will approximate the intensity experienced during the practitioner’s initial pressing and remain within the participant’s tolerance. The same verbal instructions will be used in both groups.

Participants will be instructed to keep the adhesive-covered points clean and as dry as possible during bathing and hair washing. If an adhesive becomes wet but remains attached, the participant should press it firmly back into place. Detached, contaminated, or damaged devices must not be reused or reinserted by participants. Participants will contact the study team for assessment, and any replacement by study personnel will be documented. Persistent pain, swelling, discharge, marked redness, or other local reactions should be reported promptly.

### 4.6 Practitioner Background and Standardization

Both AA and SS will be delivered by practitioners holding professional acupuncture qualifications and having 5 to 10 years of clinical acupuncture experience. All practitioners will follow the fixed-point prescription and procedures described in Sections 4.3 through 4.5. Application and treatment exposure will be documented as described in Section 4.4.

### 4.7 Comparator Rationale and Procedures

Sham units were prepared from the same commercial JIAJIAN SPN 0.22 × 2.0-mm product by cutting the protruding needle flush to form a flat contact surface that did not penetrate the skin. After trimming at the treatment site, the treating acupuncturist disinfected this surface with povidone-iodine and applied the device without repackaging. The commercial base, adhesive material, and outward appearance were unchanged. Four devices were applied using the same point sequence, treatment regimen, and participant instructions specified in Sections 4.3 through 4.5.

### 4.8 Criteria for Modifying or Discontinuing Allocated Intervention

The point prescription and device dimensions will not be individualized or adjusted. Pressure during placement and self-stimulation may be reduced to a tolerable level, but points and penetration depth will not be changed. Participants must not reinsert loose, damaged, or detached devices. Such events, and any replacement by study staff, will be documented.

The allocated intervention will be discontinued for intolerable pain, clinically important adhesive allergy, auricular infection, a serious or otherwise clinically important AE, clinical worsening or another condition that makes continued treatment unsafe, a subsequently identified eligibility error or major protocol violation, or the participant’s request. The treating clinician will remove the devices, document the decision, provide or arrange appropriate care, and report the event through the trial safety pathway. Suspected infection will require device removal, local cleansing and wound care, and topical or other anti-infective treatment when clinically indicated.

Treatment may be temporarily interrupted while a local reaction or intercurrent condition is assessed. It may resume only when judged safe and within the planned 6-week treatment period. Follow-up and analysis after intervention discontinuation will follow Section 3.7.

### 4.9 Permitted and Prohibited Concomitant Care

Medication intended to treat rosacea or reasonably expected to alter rosacea symptoms will not be used during the 1-week period before baseline or during the 6-week intervention period. This restriction includes, but is not limited to, carvedilol, gabapentin, duloxetine, minocycline hydrochloride, and isotretinoin. Any washout or discontinuation must be clinically appropriate and completed under the direction of the participant’s treating clinician; medication required for participant safety will not be withheld.

Stable treatment for unrelated medical conditions may continue if the investigator judges that it does not create a safety concern, violate an eligibility criterion, or materially affect rosacea outcomes. All prescription and nonprescription medicines and other rosacea-directed treatments used from screening through week 10 will be recorded, including the indication, dose or procedure, and start and stop dates.

## 5 Outcomes and Participant Timeline

### 5.1 Primary Outcome

The primary outcome is the CEA score at week 6, the prespecified end of the 6-week treatment period. The outcome variable is the single participant-level score recorded at that visit on the 5- category CEA scale: 0, clear (no erythema); 1, almost clear (slight redness); 2, mild (definite redness); 3, moderate (marked redness); and 4, severe (fiery redness). Lower scores indicate less severe persistent facial erythema. A dermatologist who is masked to randomized assignment will perform the live clinical assessment. The CEA has shown high inter-rater and good intra-rater reliability when used by trained raters in patients with rosacea ^16^.

CEA was selected because persistent facial erythema is the defining treatment target in this ETR population and because the scale provides a standardized, clinically interpretable assessment suitable for masked dermatologist rating. Baseline CEA is collected for covariate adjustment, as specified in Section 8. The primary treatment contrast, missing-data procedures, and supportive analysis that respects the ordinal scale are also prespecified in Section 8.

### 5.2 Secondary Outcomes

The prespecified secondary outcomes are CEA scores at weeks 2, 4, and 10 and the following scale scores at weeks 2, 4, 6, and 10. Baseline values of every scale will also be collected. Each outcome is the participant-level total or global score at the specified visit. Unless otherwise stated, lower scores indicate a more favorable clinical state. Collectively, these outcomes characterize persistent erythema, episodic flushing, migraine symptoms, sleep quality, depression, and anxiety across treatment and post-treatment follow-up.

PSA of facial redness will be reported on a 0-to-4 ordinal scale (0, no redness; 1, slight redness; 2, mild redness; 3, moderate redness; and 4, severe redness) ^17^. The PSA will be completed by the participant without access to the clinician’s CEA rating.

GFSS will capture the participant’s overall severity of facial flushing during the previous 7 days on a 0-to-10 scale, anchored at 0 (no flushing) and 10 (extremely severe flushing) ^18^. The GFSS is kept distinct from the CEA: GFSS evaluates episodic flushing, whereas CEA evaluates persistent facial erythema. This feature-specific separation is consistent with global rosacea assessment recommendations ^13^.

Migraine severity will be assessed by the participant on the study’s 0-to-10 VAS with reference to the previous 7 days, anchored at 0 (no migraine) and 10 (most severe migraine). The participant’s single recorded value will define the visit-level VAS outcome ^19^.

Sleep quality will be assessed with the PSQI, which uses a 1-month recall period and yields 7 component scores and a global score from 0 to 21. Higher scores indicate poorer sleep quality ^20^. Depression and anxiety symptom severity will be assessed with the HAMD and the HAMA, respectively, administered and scored by psychiatrists using the study versions of the instruments. Higher scores indicate greater symptom severity ^21,22^.

### 5.3 Safety and Harms Outcomes

Safety outcomes will be assessed from the first application through week 10 and will include AEs, treatment-emergent AEs, intervention-related AEs, grade 3 or higher AEs, serious adverse events (SAEs), intervention-related SAEs, temporary treatment interruptions, and permanent intervention discontinuations because of AEs. Events of particular relevance include auricular pain, local erythema or inflammation, bleeding, adhesive-related skin reactions, infection, sleep disturbance, dizziness, headache, and nausea.

Severity will be graded using the Common Terminology Criteria for Adverse Events (CTCAE) version 5.0, when an applicable term exists. Event definitions, ascertainment, documentation, and reporting procedures are specified in Section 9.3. Statistical summaries are specified in Section 8.7.

### 5.4 Outcome Measurement Properties and Standardization

Outcome definitions, instruments, scoring, assessors, and visit schedules will be standardized across all 5 sites. Masked dermatologists will receive training in the CEA grading anchors. CEA will be rated from the participant’s current clinical appearance rather than reconstructed from questionnaires or photographs. CEA and PSA ratings will be completed independently.

Participant-reported measures will be completed before treatment-related discussion at the assessment visit whenever operationally feasible. Staff may explain item wording in a neutral and standardized manner but will not coach responses. HAMD and HAMA will be administered by psychiatrists using the same item wording and scoring rules at all visits.

Facial photographs will be obtained at baseline and weeks 2, 4, 6, and 10 under each site’s standardized acquisition procedure as supporting source documentation.

The CEA, PSA, GFSS, migraine VAS, PSQI, HAMD, and HAMA rating scale are provided in Appendices A through G, respectively.

### 5.5 Participant Timeline

Potential participants will undergo screening and provide written informed consent before any trial-specific procedure. Eligibility, relevant medical history, concomitant treatments, and the required 1-week washout of prohibited rosacea-directed medication will be reviewed before baseline. At week 0, eligibility will be confirmed, baseline CEA, PSA, GFSS, VAS, PSQI, HAMD, and HAMA assessments and facial photography will be completed, and eligible participants will be randomized before the first application.

Assigned AA or SS will be applied every 3 days for 6 weeks, for 14 planned applications. Efficacy assessments and facial photography will be repeated at weeks 2 and 4 and at the end-of- treatment visit at week 6. Safety, treatment exposure, device problems, concomitant care, and protocol deviations will be reviewed throughout treatment. No study intervention will be delivered from week 6 to week 10. The final follow-up at week 10 will repeat all efficacy assessments and safety review. Participants who discontinue the intervention will be invited to complete the remaining assessments unless they withdraw consent for further data collection.

The schedule follows the SPIRIT 2013 recommendations for displaying enrollment, interventions, and assessments ^23^.

## 6 Assignment of Interventions

### 6.1 Random Sequence Generation

Eligible participants will be assigned in a 1:1 ratio to AA or SS through the central EDC system of the First Affiliated Hospital of Army Medical University. The EDC randomization module will use simple unrestricted randomization, with a probability of 0.50 for each group at each allocation. No stratification, permuted blocks, minimization, or site-specific allocation quota will be used. Exact equality of the final group sizes is therefore not guaranteed, even though the allocation probability is 1:1.

Before enrollment, data managers will configure and validate the central EDC randomization module. Thereafter, they will have no role in enrollment, allocation, outcome assessment, or statistical analysis and will not access accumulating allocations. Outcome assessors and the trial statistician will not see forthcoming assignments or be able to alter the allocation algorithm.

Assignments will be generated only at the time of randomization, with no advance lists distributed to sites.

### 6.2 Allocation Concealment

Allocation remained concealed until written informed consent had been obtained, eligibility had been confirmed, and baseline assessments had been completed. The central EDC system then generated the participant’s assignment, which was communicated to the treating acupuncturist for implementation. No advance allocation list was distributed to participating sites, and the enrolling clinician could not select, reject, override, or repeat an assignment. Central, participant- specific release preserved allocation concealment before randomization.

### 6.3 Implementation

At each site, a dermatologist identified potential participants, obtained written informed consent, confirmed eligibility, and arranged baseline assessments. After these steps, the participant was referred to the treating acupuncturist, who initiated the central randomization request and implemented the assigned intervention.

The treating acupuncturist documented the allocation date, assigned intervention, and device application in the intervention record and did not perform efficacy outcome assessment or statistical analysis.

### 6.4 Masking

Participants, dermatologists, psychiatrists, and the trial statistician will remain masked to allocation until database lock and code release. Treating acupuncturists cannot be masked because they must apply the allocated device, but they will not assess efficacy outcomes or conduct statistical analyses.

Masking will be supported by the similar appearance and adhesive of AA and sham devices. The active needle will penetrate the skin, whereas the sham device will not. Point locations, skin preparation, visit schedule, retention, ear alternation, self-pressing instructions, and practitioner contact will be matched. Participants will not be shown the devices during placement and will be instructed not to discuss device sensations with outcome assessors.

### 6.5 Permissible Unmasking and Emergency Procedures

Because treating acupuncturists are already aware of assignment, most immediate intervention actions—including removal of the device, temporary interruption, or permanent discontinuation—can be taken without revealing assignment to masked personnel. Individual unmasking will be permitted only when knowledge of the assigned device is necessary for the urgent clinical management of a serious or otherwise clinically important AE and the required information cannot be obtained adequately from the unmasked treating acupuncturist. Suspected lack of efficacy, routine AE reporting, protocol deviation, withdrawal, or administrative convenience will not justify unmasking.

If emergency unmasking is needed, a delegated physician will request an authorized participant- specific query in the EDC system. The site will record the participant identifier, date, time, clinical justification, and actions taken. The coordinating center and, when required, the ethics committee will be notified promptly. Staff not involved in the emergency will remain masked when possible. Unmasking alone will not end study participation, and scheduled follow-up will continue unless consent is withdrawn or further participation is clinically inappropriate.

## 7 Data Collection and Management

### 7.1 Data Collection Methods and Instruments

Trial data will be collected at screening, baseline (week 0), weeks 2, 4, 6, and 10, and the treatment contacts scheduled every 3 days during the 6-week intervention, as applicable to each measure and shown in the Schedule of Assessments. Data will first be recorded in contemporaneous source documents and paper CRFs and will then be entered promptly into the central EDC system. The records will cover consent and eligibility, demographic and clinical characteristics, outcome assessments, intervention exposure and fidelity, concomitant care, protocol deviations, withdrawals, and AEs.

At baseline and scheduled outcome visits, a masked dermatologist will assess CEA. Participants will complete the PSA, GFSS, migraine VAS, and PSQI. Psychiatrists will administer the HAMD and HAMA. Adverse events, device problems, treatment exposure, concomitant care, and withdrawal or deviation status will be reviewed at the specified visits (Sections 4 and 5).

Standardized facial photographs will be obtained at baseline and weeks 2, 4, 6, and 10 as supporting source documentation and will not replace the contemporaneous masked CEA rating. No photographs of the auricular intervention sites were obtained.

### 7.2 Assessor Procedures and Measurement Standardization

Dermatologists will receive study-specific training on the 5 CEA anchors and will score erythema without referring to PSA responses or treatment records. Two dermatologists will independently score baseline CEA before allocation. If they disagree, they will review the participant together and agree on a consensus baseline score before randomization. Both raters must confirm a CEA score of at least 2 for eligibility. Follow-up CEA scores will be obtained by a masked dermatologist.

Staff administering the PSA, GFSS, migraine VAS, PSQI, HAMD, and HAMA will use the same item wording, recall periods, response anchors, and scoring rules at all sites.

### 7.3 Participant Retention and Follow-up

The visit schedule and the importance of completing outcome assessments will be explained during consent and reinforced at treatment contacts. With the participant’s permission, site staff will provide visit reminders by telephone or WeChat. Contact details and preferred permissible contact methods will be confirmed during follow-up.

If a participant misses a visit and has not withdrawn consent, site staff will make reasonable, documented attempts by the permitted contact methods to re-establish contact and complete the remaining assessments. The date of last contact, attempts made, follow-up status, and reason for noncompletion, when known, will be recorded.

Protocol deviations or clinically required concomitant care will not automatically end data collection. Follow-up after intervention discontinuation or withdrawal of consent will follow Section 3.7.

### 7.4 Data Entry, Coding, and Quality Assurance

Data managers will manually review records for completeness, permissible values, consistency, visit timing, and agreement with source information. Before database lock, a final review will address participant disposition, outcomes, treatment exposure, adverse events, deviations, and unresolved queries. Outcome assessors and the trial statistician will remain masked until database lock. Treatment codes will be released for the final analysis only after lock.

## 8 Statistical Methods

### 8.1 Statistical Objectives and Estimands

This 2-group superiority trial will estimate the effect of assignment to AA versus SS on CEA at week 6. In the ICH E9(R1) estimand framework, the target population is all randomized participants, and the treatment conditions are assignment to AA or SS with the common study procedures. The outcome variable is the week 6 participant-level CEA score, summarized as the baseline-adjusted mean difference (AA minus SS). Treatment discontinuation, missed applications, nonadherence, and clinically required concomitant or rescue treatment will be handled using a treatment-policy strategy. A negative difference favors AA because lower CEA scores reflect less severe erythema.

Participants will be analyzed according to their assigned group, and observed outcomes collected after intercurrent events will remain relevant to the primary estimand. Missing week 6 CEA observations will be handled using the planned missing-data methods rather than by changing the analysis population. Secondary outcomes will use the same treatment-policy strategy at their scheduled visits. Safety will be summarized among participants receiving at least 1 intervention.

### 8.2 Sample Size Determination

The sample size was calculated based on the between-group difference in the week 6 CEA score. A mean difference of 0.70 points was considered a clinically relevant treatment effect, with a common SD of 1.20. With a 2-sided alpha level of .05, 80% power, and a 1:1 allocation ratio, 48 participants per group were required. The calculation was performed using PASS 2023 software (NCSS, LLC).

Allowing for 10% attrition, the planned sample size was increased to 54 participants per group, for a total of 108 participants.

### 8.3 Analysis Populations

The ITT population will include every randomized participant, analyzed in the group assigned at randomization. The ITT population is the primary population for all efficacy analyses and includes participants who receive no intervention, discontinue treatment, miss applications, use concomitant or rescue treatment, or have a protocol deviation. A participant with no observed week 6 CEA score will remain in the ITT population and will be represented in the primary analysis through the prespecified missing-data procedure.

The safety population will include all randomized participants who receive at least 1 application of the trial intervention and will be analyzed according to the intervention actually received. A complete-case population will include participants in the ITT population with the observed baseline and week 6 CEA values required by the ANCOVA and will be used only for sensitivity analysis.

### 8.4 General Analysis Principles

Treatment groups will be identified by masked codes during data review and programming. Analyses will be conducted after database lock with 2-sided tests. For the primary comparison, the significance level will be .05, with a 95% CI. Interpretation will emphasize effect estimates and CIs rather than P values alone. All contrasts will be expressed as AA minus SS, with the direction of benefit specified for each scale.

Continuous variables will be summarized using means and SDs and, when useful for skewed distributions, medians and interquartile ranges. Categorical variables will be summarized using counts and percentages with explicit denominators. Baseline characteristics will be described by randomized group without significance testing, because any baseline differences arise after randomization and do not require a test of the randomization process. Source-verified extreme observations will not be removed solely because they affect normality tests. The primary model will adjust for baseline CEA and randomized group.

### 8.5 Primary Outcome Analysis

Week 6 CEA will be analyzed using ANCOVA with randomized group and baseline CEA as model terms. The treatment effect will be reported as the adjusted mean difference (AA minus SS), with a 95% CI and 2-sided P value. Adjusted group means will also be reported. Missing data will be handled as specified in Section 8.9.

Model diagnostics will assess linearity with baseline CEA, residual variance, residual normality, and influential observations. Because CEA has 5 ordered categories, an adjusted proportional- odds ordinal logistic model will be used for support. The common odds ratio will describe the odds of a higher (worse) CEA category for AA versus SS. Values below 1 favor AA.

### 8.6 Secondary and Exploratory Outcome Analyses

Repeated CEA scores and the PSA, GFSS, migraine VAS, PSQI, HAMD, and HAMA scores will be analyzed using separate mixed models for repeated measures. For each outcome, the postbaseline score will be the dependent variable and the fixed terms will be randomized group, categorical visit, the group-by-visit interaction, the corresponding baseline score, and the baseline-by-visit interaction. Within-participant correlation will be modeled using an unstructured residual covariance matrix with visit-specific residual variances. Model-based adjusted means and adjusted mean differences (AA minus SS) with 95% CIs will be estimated at each scheduled visit.

The repeated-measures models will use all available postbaseline observations under a MAR assumption and will not use single imputation. CEA contrasts at weeks 2, 4, and 10 and all PSA, GFSS, migraine VAS, PSQI, HAMD, and HAMA contrasts at weeks 2, 4, 6, and 10 are secondary.

### 8.7 Analysis of Harms

Harms will be summarized in the safety population according to treatment actually received. AEs will be graded with the CTCAE version 5.0, when an applicable term exists. For each group, the number and percentage of participants with at least 1 AE, treatment-emergent AE, intervention- related AE, grade 3 or higher event, SAE, intervention-related SAE, temporary treatment interruption, and permanent discontinuation because of an event will be reported.

Events will be summarized by their recorded adverse event names, maximum CTCAE version 5.0 grade, seriousness, investigator-assessed relatedness, action taken, and outcome. Both the number of affected participants and the number of events will be shown, and the denominator for percentages will be stated. Treatment-emergent events through week 6 will be distinguished from events first identified during follow-up through week 10.

### 8.8 Multiplicity

There is 1 confirmatory primary comparison: AA vs SS for the week 6 CEA score. Its 2-sided type I error rate will be controlled at 0.05, and no multiplicity adjustment is required for that single test. All comparisons for CEA at other visits and for PSA, GFSS, migraine VAS, PSQI, HAMD, and HAMA are secondary and exploratory in interpretation. Their P values and 95% CIs will be nominal and will not be adjusted for multiple outcomes or visits.

### 8.9 Missing Data

The amount, timing, pattern, and documented reasons for missing outcome data will be summarized by randomized group and visit. For the primary analysis, missing week 6 CEA scores will be handled by MI under a MAR assumption. Fifty imputed data sets will be created using predictive mean matching. The imputation model will include randomized group; baseline and all available postbaseline CEA scores; and baseline PSA, GFSS, migraine VAS, PSQI, HAMD, and HAMA scores. The ANCOVA will be fitted in each imputed data set and the estimates and standard errors combined using Rubin’s rules. The likelihood-based repeated- measures analyses will use all observed values without separate imputation under their MAR assumption.

### 8.10 Sensitivity Analyses

Robustness of the primary conclusion will be evaluated using a complete-case ANCOVA with randomized group and baseline CEA, the baseline-adjusted proportional-odds model described above, and an extreme missing-data analysis. The extreme analysis will assign a CEA score of 4 to every missing AA week 6 outcome and a score of 0 to every missing SS week 6 outcome before fitting the baseline-adjusted ANCOVA.

### 8.11 Statistical Software and Reproducibility

Analyses will be conducted in R version 4.4.0. Multiple imputation will use mice version 3.16.0, and repeated-measures models will use nlme version 3.1-165. Package versions, R session information, model formulas, contrast direction, and random-number seeds used for imputation will be retained with the analysis programs.

The trial statistician will conduct the analyses from a read-only extract of the locked database after release of the treatment codes. Derived variables and analysis populations will be generated programmatically where feasible. The analysis code, execution log, model diagnostics, imputation diagnostics, and final tables and figures will be archived in the statistical trial file so that each reported estimate can be traced to the locked data extract.

## 9 Monitoring and Safety Oversight

### 9.1 Data Monitoring Committee (DMC)

No independent DMC will be established. The trial evaluates a short-duration, minimally invasive outpatient intervention in 108 participants, and the foreseeable intervention-related risks are predominantly local, promptly detectable, and clinically manageable. These features support direct safety oversight by the investigator team rather than a separate committee.

### 9.2 Interim Analyses and Stopping Guidelines

No interim analysis of efficacy, futility, or between-group safety outcomes is planned, and no accumulating comparative outcome results will be reviewed before database lock and unblinding. Consequently, no statistical alpha-spending function or efficacy or futility boundary is specified. Recruitment and follow-up will continue to the planned sample size unless a safety, ethical, regulatory, or trial-integrity concern requires suspension or termination.

### 9.3 AE and SAE Procedures

Safety surveillance will begin with the first application and continue through week 10. At each treatment contact and visit, adverse events will be identified through participant reports, nonleading questions, and clinical assessment. An adverse event is any unfavorable medical occurrence after treatment, regardless of relatedness. Treatment-emergent events are those that occur or worsen from the first application through week 6. Events first identified during weeks 6 to 10 will be distinguished in the safety records.

A SAE is an event that results in death, is life-threatening, requires or prolongs inpatient hospitalization, results in persistent or clinically important disability or incapacity, results in a congenital anomaly or birth defect, or is another important medical event that may jeopardize the participant or require intervention to prevent one of these outcomes. Seriousness will be assessed separately from severity. Severity will be graded using the CTCAE version 5.0, when an applicable term exists.

For each event, investigators will record its verbatim description, onset and resolution dates, severity, seriousness, treatment action, care provided, outcome, and assessed relatedness.

Relevant events include auricular pain, local inflammation, bleeding, adhesive reactions, infection, sleep disturbance, dizziness, headache, and nausea. Unanticipated events will also be reported.

The treating clinician will provide or arrange appropriate care and may remove the device, interrupt treatment, or permanently discontinue the intervention when clinically indicated. Suspected auricular infection requires device removal, local cleansing and wound care, and topical or systemic anti-infective treatment when clinically indicated. Emergency unmasking is permitted only when knowledge of the assigned device is necessary for urgent clinical management and cannot be obtained adequately from the unmasked treating practitioner.

A site investigator must report every SAE to the trial safety lead within 24 hours after becoming aware of the event, using the information then available. A complete follow-up report will be provided as additional clinical information becomes available. AEs will be followed until resolution, return to baseline, stabilization, establishment of a chronic condition, determination that further follow-up is not clinically indicated, loss to follow-up despite reasonable attempts, or death.

## 10 Ethics and Participant Protection

### 10.1 Research Ethics Approval

The trial was conducted in accordance with the ethical principles of the World Medical Association Declaration of Helsinki and applicable requirements in China, including the Measures for Ethical Review of Life Science and Medical Research Involving Human Participants.

The protocol and informed consent form will be reviewed and approved by the institutional review boards of the First Affiliated Hospital of Army Medical University and each participating center before enrollment begins at the respective center. Each center must retain its ethics approval documentation and implement the approved consent, intervention, safety, and data- protection procedures. The principal investigator is responsible for maintaining ethics approval and responding to requests from the relevant institutional review boards throughout the trial.

Eligible participants must be adults capable of providing valid informed consent. Written consent will be obtained by an authorized investigator or delegate before any study-specific procedure.

Refusal or subsequent withdrawal will not affect routine care, legal rights, or access to indicated treatment.

### 10.2 Privacy and Confidentiality

Participant confidentiality will be protected before enrollment, throughout follow-up, during analysis and publication, and throughout the record-retention period. Each participant will be identified in the EDC system and analysis files by a unique trial identifier. The site identification record and signed consent form will be stored separately from coded research data. Direct identifiers will not be included in the analysis data set or aggregate trial reports. Authorized ethics, institutional, or regulatory reviewers may inspect identifiable source records when required, subject to confidentiality obligations.

Facial photographs will be handled as potentially identifiable records and used only within this trial. No identifiable participant photograph will be published. Electronic data and photographs will be stored on the internal server of the First Affiliated Hospital of Army Medical University, and paper consent forms, CRFs, and other source documents will be kept in locked cabinets at the participating sites. Records will be retained for 5 years after trial completion, or longer if required by an ethics committee, institution, or applicable law.

The research team will use the minimum participant information necessary to conduct the trial and answer the prespecified research questions. Individual participant data and trial photographs will not be provided to external investigators for secondary research under the present consent. At the end of the required retention period, the records will be disposed of or retained further only in accordance with institutional, ethical, and legal requirements.

## Declarations

**Funding:** Internal institutional grants 202411TZDB09 from The First Affiliated Hospital of Army Medical University (see Section 11).

**Ethics approval:** Study procedures were reviewed and approved by the ethics committee of the First Affiliated Hospital of Army Medical University ((A) KY2024102).

**Competing interests:** All authors declare no conflicts of interest.

**Data availability:** Participant-level data and facial photographs are not available for external secondary research under the consent described in Section 10.2.

## List of Abbreviations and Definitions

### Abbreviation or term Definition

AA: Auricular Acupuncture
AE: Adverse Event
ANCOVA: Analysis of Covariance
CEA: Clinician Erythema Assessment
CRF: Case Report Form
CTCAE: Common Terminology Criteria for Adverse Events
DMC: Data Monitoring Committee
EDC: Electronic Data Capture
ETR: Erythematotelangiectatic Rosacea
GFSS: Global Flushing Severity Score
HAMA: Hamilton Anxiety Rating Scale
HAMD: Hamilton Depression Rating Scale
ITT: Intention-to-Treat
MAR: Missing at Random
PSA: Patient Self-Assessment
PSQI: Pittsburgh Sleep Quality Index
SAE: Serious Adverse Event
SAP: Statistical Analysis Plan
SPN: Sterile Press Needle
SS: Sham Stimulation
VAS: Visual Analog Scale

## Data Availability

Participant-level data and facial photographs are not available for external secondary research under the consent.

## 12 Outcome Instrument Appendices

### Appendix A Clinician Erythema Assessment

| **Grade** | **Description** |
|---|---|
| 0 | Clear skin with no signs of erythema |
| 1 | Almost clear; slight redness |
| 2 | Mild erythema; definite redness |
| 3 | Moderate erythema; marked redness |
| 4 | Severe erythema; fiery redness |

### Appendix B Patient Self-Assessment

| **Score** | **Description** |
|---|---|
| 0 | No redness |
| 1 | Slight redness |
| 2 | Mild redness |
| 3 | Moderate redness |
| 4 | Severe redness |

### Appendix C Global Flushing Severity Score

| **Score** | **Severity descriptor** | **Selected score** |
|---|---|---|
| 0 | None | [ ] |
| 1-3 | Mild | [ ] |
| 4-6 | Moderate | [ ] |
| 7-9 | Severe | [ ] |
| 10 | Extreme | [ ] |
| Recall period used in this trial | Overall facial flushing severity during the previous 7 days | ____ / 10 |

### Appendix D Visual Analog Scale

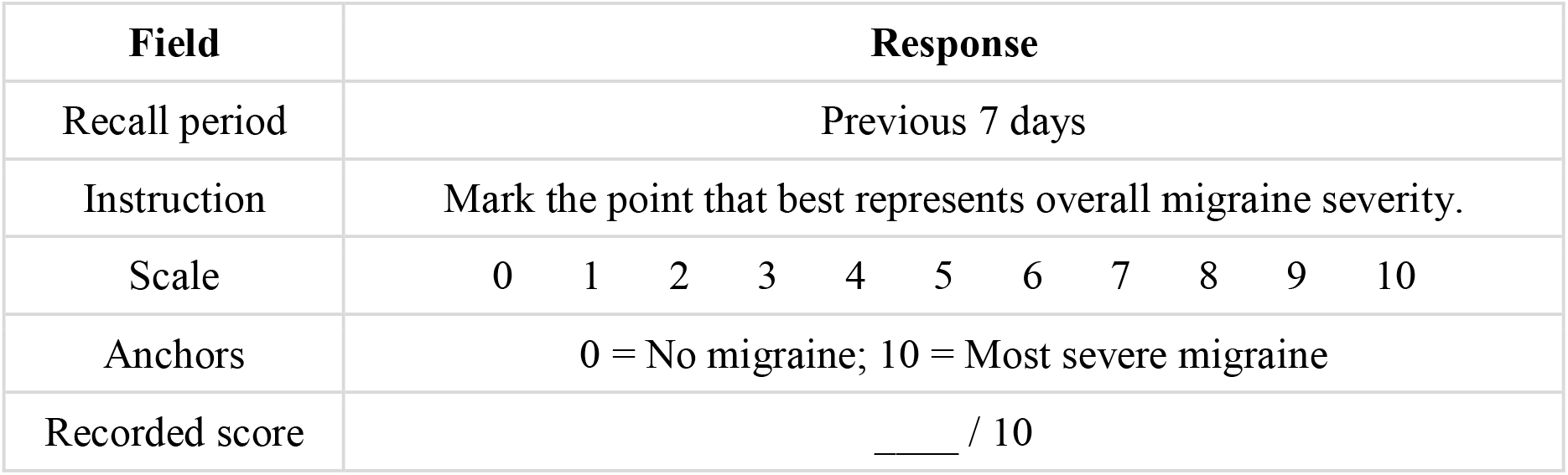

| **Field** | **Response** |
|---|---|
| Recall period | Previous 7 days |
| Instruction | Mark the point that best represents overall migraine severity. |
| Scale | 0 1 2 3 4 5 6 7 8 9 10 |
| Anchors | 0 = No migraine; 10 = Most severe migraine |
| Recorded score | ____ / 10 |

### Appendix E Pittsburgh Sleep Quality Index

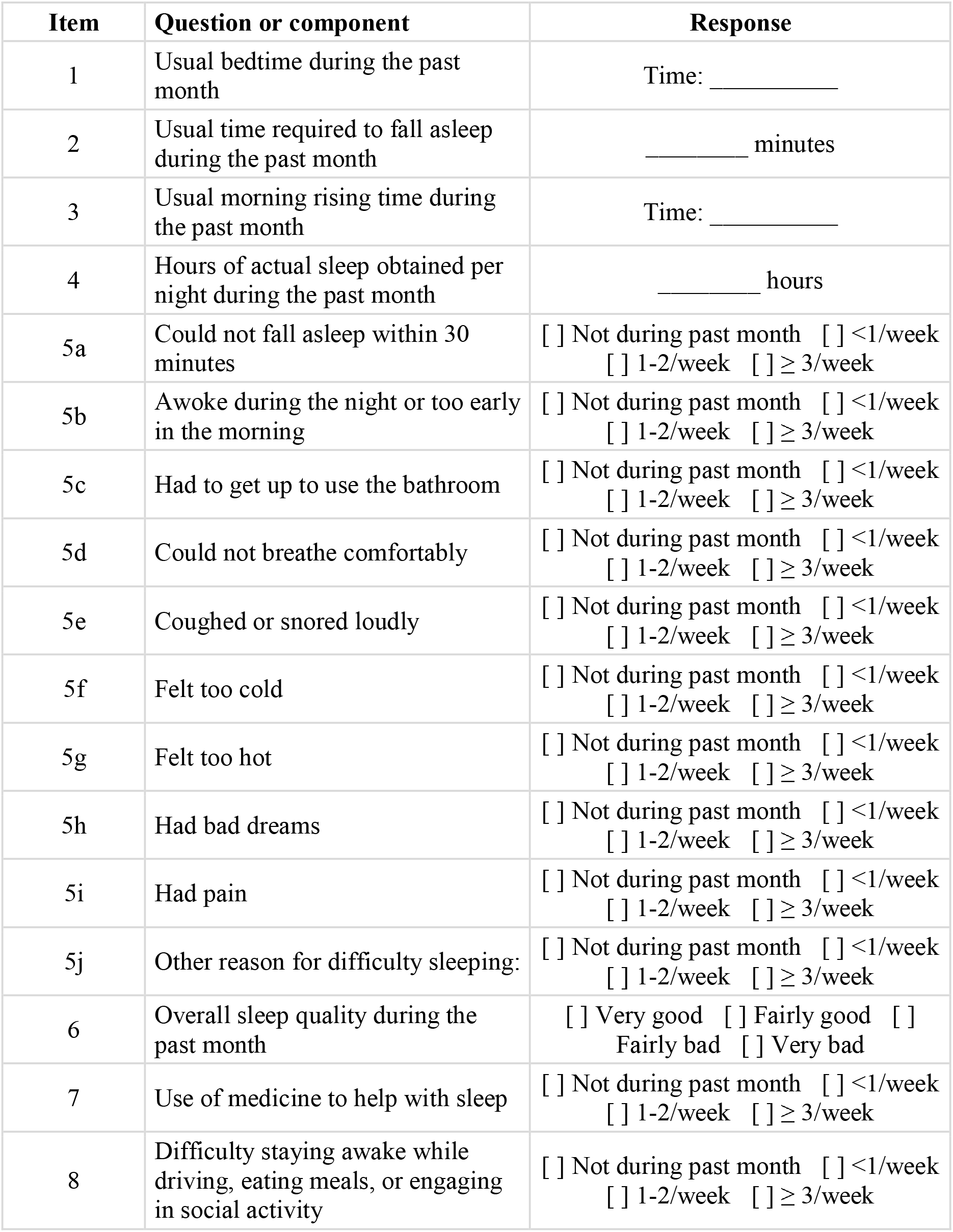
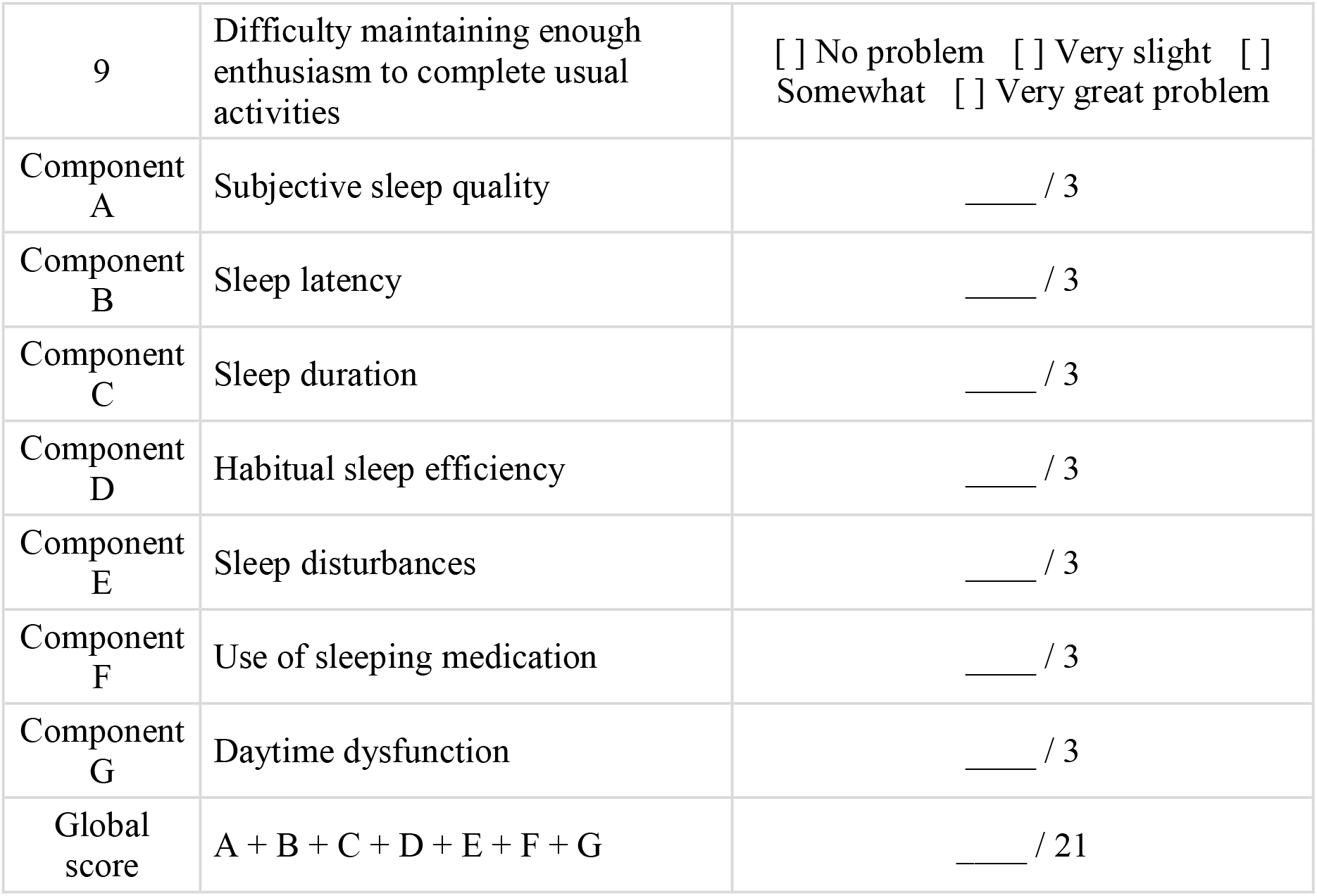

| Item | Question or component | Response |
|---|---|---|
| 1 | Usual bedtime during the past month | Time: __________ |
| 2 | Usual time required to fall asleep during the past month | ________ minutes |
| 3 | Usual morning rising time during the past month | Time: __________ |
| 4 | Hours of actual sleep obtained per night during the past month | ________ hours |
| 5a | Could not fall asleep within 30 minutes | [ ] Not during past month [ ] <1/week [ ] 1-2/week [ ] ≥ 3/week |
| 5b | Awoke during the night or too early in the morning | [ ] Not during past month [ ] <1/week [ ] 1-2/week [ ] ≥ 3/week |
| 5c | Had to get up to use the bathroom | [ ] Not during past month [ ] <1/week [ ] 1-2/week [ ] ≥ 3/week |
| 5d | Could not breathe comfortably | [ ] Not during past month [ ] <1/week [ ] 1-2/week [ ] ≥ 3/week |
| 5e | Coughed or snored loudly | [ ] Not during past month [ ] <1/week [ ] 1-2/week [ ] ≥ 3/week |
| 5f | Felt too cold | [ ] Not during past month [ ] <1/week [ ] 1-2/week [ ] ≥ 3/week |
| 5g | Felt too hot | [ ] Not during past month [ ] <1/week [ ] 1-2/week [ ] ≥ 3/week |
| 5h | Had bad dreams | [ ] Not during past month [ ] <1/week [ ] 1-2/week [ ] ≥ 3/week |
| 5i | Had pain | [ ] Not during past month [ ] <1/week [ ] 1-2/week [ ] ≥ 3/week |
| 5j | Other reason for difficulty sleeping: | [ ] Not during past month [ ] <1/week [ ] 1-2/week [ ] ≥ 3/week |
| 6 | Overall sleep quality during the past month | [ ] Very good [ ] Fairly good [ ] Fairly bad [ ] Very bad |
| 7 | Use of medicine to help with sleep | [ ] Not during past month [ ] <1/week [ ] 1-2/week [ ] ≥ 3/week |
| 8 | Difficulty staying awake while driving, eating meals, or engaging in social activity | [ ] Not during past month [ ] <1/week [ ] 1-2/week [ ] ≥ 3/week |
| 9 | Difficulty maintaining enough enthusiasm to complete usual activities | [ ] No problem [ ] Very slight [ ] Somewhat [ ] Very great problem |
| Component A | Subjective sleep quality | ____ / 3 |
| Component B | Sleep latency | ____ / 3 |
| Component C | Sleep duration | ____ / 3 |
| Component D | Habitual sleep efficiency | ____ / 3 |
| Component E | Sleep disturbances | ____ / 3 |
| Component F | Use of sleeping medication | ____ / 3 |
| Component G | Daytime dysfunction | ____ / 3 |
| Global score | A + B + C + D + E + F + G | ____ / 21 |

### Appendix F Hamilton Depression Rating Scale

| **No.** | **HAMD item** | **0** | **1** | **2** | **3** | **4** |
|---|---|---|---|---|---|---|
| 1 | Depressed mood | [ ] | [ ] | [ ] | [ ] | [ ] |
| 2 | Feelings of guilt | [ ] | [ ] | [ ] | [ ] | [ ] |
| 3 | Suicide | [ ] | [ ] | [ ] | [ ] | [ ] |
| 4 | Insomnia, early | [ ] | [ ] | [ ] | - | - |
| 5 | Insomnia, middle | [ ] | [ ] | [ ] | - | - |
| 6 | Insomnia, late | [ ] | [ ] | [ ] | - | - |
| 7 | Work and activities | [ ] | [ ] | [ ] | [ ] | [ ] |
| 8 | Psychomotor retardation | [ ] | [ ] | [ ] | [ ] | [ ] |
| 9 | Agitation | [ ] | [ ] | [ ] | [ ] | [ ] |
| 10 | Psychic anxiety | [ ] | [ ] | [ ] | [ ] | [ ] |
| 11 | Somatic anxiety | [ ] | [ ] | [ ] | [ ] | [ ] |
| 12 | Somatic symptoms, gastrointestinal | [ ] | [ ] | [ ] | - | - |
| 13 | Somatic symptoms, general | [ ] | [ ] | [ ] | - | - |
| 14 | Genital symptoms | [ ] | [ ] | [ ] | - | - |
| 15 | Hypochondriasis | [ ] | [ ] | [ ] | [ ] | [ ] |
| 16 | Loss of weight | [ ] | [ ] | [ ] | - | - |
| 17 | Insight | [ ] | [ ] | [ ] | - | - |
| 18 | Diurnal variation | [ ] | [ ] | [ ] | - | - |
| 19 | Depersonalization and derealization | [ ] | [ ] | [ ] | [ ] | [ ] |
| 20 | Paranoid symptoms | [ ] | [ ] | [ ] | [ ] | [ ] |
| 21 | Obsessional and compulsive symptoms | [ ] | [ ] | [ ] | - | - |
| 22 | Helplessness | [ ] | [ ] | [ ] | [ ] | [ ] |
| 23 | Hopelessness | [ ] | [ ] | [ ] | [ ] | [ ] |
| 24 | Worthlessness | [ ] | [ ] | [ ] | [ ] | [ ] |
|  | Total score |  |  |  |  |  |

### Appendix G Hamilton Anxiety Rating Scale

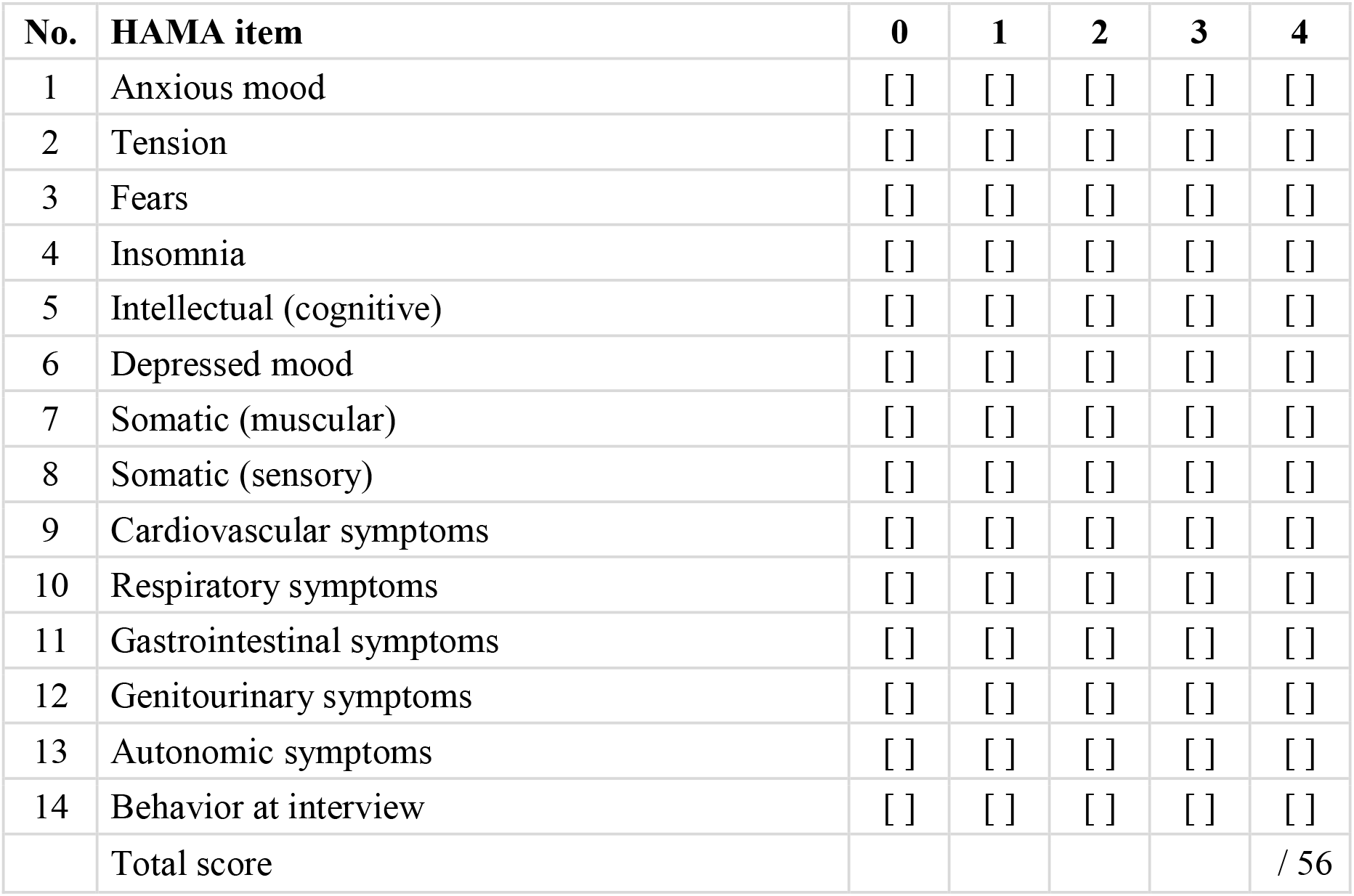

| **No.** | **HAMA item** | **0** | **1** | **2** | **3** | **4** |
|---|---|---|---|---|---|---|
| 1 | Anxious mood | [ ] | [ ] | [ ] | [ ] | [ ] |
| 2 | Tension | [ ] | [ ] | [ ] | [ ] | [ ] |
| 3 | Fears | [ ] | [ ] | [ ] | [ ] | [ ] |
| 4 | Insomnia | [ ] | [ ] | [ ] | [ ] | [ ] |
| 5 | Intellectual (cognitive) | [ ] | [ ] | [ ] | [ ] | [ ] |
| 6 | Depressed mood | [ ] | [ ] | [ ] | [ ] | [ ] |
| 7 | Somatic (muscular) | [ ] | [ ] | [ ] | [ ] | [ ] |
| 8 | Somatic (sensory) | [ ] | [ ] | [ ] | [ ] | [ ] |
| 9 | Cardiovascular symptoms | [ ] | [ ] | [ ] | [ ] | [ ] |
| 10 | Respiratory symptoms | [ ] | [ ] | [ ] | [ ] | [ ] |
| 11 | Gastrointestinal symptoms | [ ] | [ ] | [ ] | [ ] | [ ] |
| 12 | Genitourinary symptoms | [ ] | [ ] | [ ] | [ ] | [ ] |
| 13 | Autonomic symptoms | [ ] | [ ] | [ ] | [ ] | [ ] |
| 14 | Behavior at interview | [ ] | [ ] | [ ] | [ ] | [ ] |
|  | Total score |  |  |  |  | / 56 |

